# Putative breast cancer risk variants from populations of South Asian ancestry are under-represented in public variant classification databases

**DOI:** 10.1101/2025.02.13.25322245

**Authors:** Raveen Rony, Shenglong Deng, Sarah Yang, Ken Doig, David L Goode

## Abstract

The majority of publicly available genomics data originates from populations of European ancestry. This limits understanding and detection of inherited genetic risk factors for breast cancer in other populations. To assess the extent to which deficits in knowledge of the genetics of breast cancer risk exist for populations of non-European ancestry, we compared data available on putative breast cancer risk variants in the ClinVar database for populations of different ancestry.

Protein-coding insertions and deletions (indels) and single-nucleotide polymorphisms (SNPs) private to populations of Non-Finnish European (NFE), African (AFR), Admixed American (AMR), East Asian (EAS) and South Asian (SAS) ancestry from the Genome Aggregation Consortium (gnomAD v4) were identified for nine established breast cancer risk genes. The percentage of private protein-coding variants listed as ‘Unreported’ by gnomAD in ClinVar were compared between populations.

The SAS population had the biggest knowledge deficit, as 43.4% of private SAS variants were not reported in ClinVar, compared to 20-30% for other populations. Proportionally fewer SAS variants were reported for all 9 genes, with the difference reaching an adjusted p < 0.05 for PALB2, ATM and BRCA2 when compared to NFE. In contrast, few genes had significantly lower ClinVar reporting rates for AFR, AMR and EAS than for NFE.

ClinVar reporting deficits in the SAS population were observed for both missense and protein-truncating variants. Unreported variants were usually very rare and largely absent in other public repositories. A substantial fraction of unreported variants were protein-truncating (17.2%), or missense with high predicted pathogenicity scores, representing novel candidate breast cancer risk alleles.

Our work demonstrates putative breast cancer risk variants from populations of South Asian ancestry are less likely to be reported in ClinVar. Defining and removing barriers to reporting potential risk variants for breast cancer from South Asian populations is needed to reduce this knowledge deficit.

## Introduction

Most cancer research to date has been conducted with patients with European ancestry (AACR CANCER DISPARITIES PROGRESS REPORT 2024, 2024) leading to deficits in our knowledge about cancer in populations with other ancestries (Aldrighetti et al., 2021; Molina-Aguilar & Robles-Espinoza, 2023) that contribute to inequities in quality of care and treatment outcomes in patients of non-European ancestry (Daly & Olopade, 2015; Garraway et al., 2024). Recent work shows the genomics and morphology of cancer can vary greatly between patients of European, African and Asian ancestries (Huang et al., 2017; Lang et al., 2020; Le et al., 2023). This impacts efforts to personalize patient treatments based on specific clinical and molecular tumour features (Edsjö et al., 2023). If precision oncology reflects existing biases towards patients of European ancestry (Aldrighetti et al., 2021; Le et al., 2023), they will only sustain existing inequities in cancer care (Arora et al., 2022).

Personalized approaches based on patient genomics have been incorporated clinically to assess individual risk of breast cancer (Yadav & Couch, 2019). Identification of putative inherited risk variants via germline DNA sequencing enables women and their families to implement individualized risk-reduction strategies (Tischler et al., 2019). However, the comparative analyses of public data on rare variants with potential to increase breast cancer risk across populations of different ancestry are lacking. ClinVar (Landrum et al., 2020) aggregates expert report evaluating links between genetic variants and human disease, including breast cancer, and is the most comprehensive and prominent database of its type. We reasoned gaps in knowledge about risk factors for breast cancer in non-European populations would result in them having proportionally fewer reports in ClinVar on the pathogenicity of variants in breast cancer associated genes.

Our study focused on nine genes (*ATM, BRCA1, BRCA2, CHEK2, PALB2, RAD51C, RAD51D, TP53*) where the Breast Cancer Association Consortium (BCAC) found inherited protein-truncating variants to be associated with elevated risk of developing breast cancer (Breast Cancer Association Consortium, 2021). Protein-coding single nucleotide polymorphisms (SNPs), insertions and deletions (indels) from the nine BCAC genes for private variants unique to one of the five populations were extracted from the Genome Aggregation Database (gnomAD) (Karczewski et al., 2020): Non-Finnish European (NFE), African (AFR), Admixed American (AMR), East Asian (EAS) and South Asian (SAS). The proportion of such population private variants reported in ClinVar for each gene and population were used to compare the extent of publicly available data on putative breast cancer risk variants by ancestry.

Our study revealed proportionally fewer expert reports of putative breast cancer risk variants from the SAS population are available in ClinVar for 7/9 genes. In contrast, ClinVar reporting rates for private variants from AFR, AMR and EAS were largely comparable with NFE across all genes, with few exceptions. The existence of such knowledge deficits advocate for more extensive assessment and reporting of putative breast cancer risk variants from individuals of South Asian ancestry.

## Methods

### Extraction and filtering of data from gnomAD

Variant data were downloaded from gnomAD v4.0.0 on March 12, 2024. The tidyverse v2.0.0 package as implemented for R software version 4.3.3 was applied to extract variants present exclusively within (i.e., private to) the Non-Finnish European (NFE), African American (AFR), Admixed American (AMR), East Asian (EAS), and South Asian (SAS) populations. Private variants were identified as those having an allele count >=1 in a population and allele count ==0 in every other population included in gnomAD v4. Population ancestry was assigned by the gnomAD consortium based on ancestry informative markers [REF]. Data were downloaded (Sept 4, 2023) and filtered in the same manner from gnomAD v2.1.0.

ClinVar status as listed in gnomAD v4.0.0 were simplified into four groups: Pathogenic, Benign, Uncertain, or Unreported. The Pathogenic category included variants labeled as Pathogenic, Likely Pathogenic, or Pathogenic/Likely Pathogenic, with a similar approach applied to the Benign variants. Annotations were assigned by gnomAD consortium, based on Variant Effect Predictor v105 using GENCODE v39 on GRCh38, with the LOFTEE (Loss-Of-Function Transcript Effect Estimator) plugin (Karczewski et al., 2020).

Fisher’s Exact test as implemented in the fisher.test() function of R v4.3.3 was applied to assess statistical significance of the difference in proportion of Unreported and Pathogenic private variants between the Non-Finnish European (NFE) population and the other populations.

### LiftOver Process

Conversion of chromosomal positions of population-private variants from gnomAD v4.0.0, based on coordinates from Genome Reference Consortium Human Build 38 (GRCh38 or hg38), to genome positions for gnomAD v2.1.0, based on coordinates from Genome Reference Consortium Human Build 37 (GRCh37 or hg19), was performed using chain file “hg38ToHg19.over.chain.gz” downloaded from https://hgdownload.soe.ucsc.edu/goldenPath/hg38/liftOver/. Coordinate conversion was implemented using the rtracklayer v1.64.0 and GenomicRanges v1.56.0 packages in RStudio.

### Retrieval and mapping of variants to other databases

The oncogenic status of Unreported population private variants from gnomAD v4.0.0 was obtained from the OncoKB database on April 16 2024 after conversion to hg19 positions for cross-referencing. The FLOSSIES database was searched on April 30, 2024 for presence of variants from gnomAD v4.0.0 with Unreported status in ClinVar.

### Prediction and scoring of population private variants using dbNSFP

Population-private variants underwent comprehensive predictive analysis and scoring using algorithms embedded within the dbNSFP v4.7a database, accessed June 5 2024. Specifically, predictions from AlphaMissense, FATHMM, MetaLR, MetaSVM, MutationAssessor, Polyphen HDIV, and SIFT, along with rankscores from AlphaMissense, CADD raw, CADD hg19, FATHMM, GERP++, MetaLR, MetaSVM, MutationAssessor, Polyphen HDIV, Polyphen HVAR, REVEL, SIFT, and VEST4. Only algorithms with rankscores between 0-1 were included for further analysis. The number of rankscores available in dbNSFP for the selected algorithms varied between genes and variants.

### Principal Component Analysis

Principal component analysis (PCA) as implemented in the prcomp() function of R v4.3.3 was used to cluster variants based on rankscores. PCA was based on 24 rankscores in common across all genes (Supp Table 11). Only rankscores with values between 0-1 were included in the PCA, to remove confounding effects of differences in ranges of values reported by different algorithms. PCA results were visualized with marginal boxplots of the distribution of each ClinVar clinical status, which was added using ggExtra.

### Code Availability

Code and intermediate data files used in these analyses are available as Git repo https://github.com/RaveenRony/ClinVar-annotation-rates)

## Results

### Identification and characterization of putative population-specific breast cancer risk variants

To compare putative breast cancer risk variants between populations, we analyzed insertion/deletion (indel) and single nucleotide variants (SNVs) from 9 nine genes reported as enriched for protein-truncating variants in breast cancer patients by the Breast Cancer Association Consortium (BCAC) in 2021: *ATM, BRCA1, BRCA2, CHEK2, PALB2, BARD1, RAD51C, RAD51D*, and *TP53*. BCAC additionally reported five of these genes (*ATM, BRCA1, BRCA2, CHEK2* and *TP53)* to be enriched for rare missense variants as well (Breast Cancer Association Consortium, 2021).

Data on variants predicted to alter protein coding sequence (frameshift indels, inframe indels, missense, splice site, stop gain or lost start) were downloaded from gnomAD v4.0 (Methods) for each of the 9 risk genes identified by BCAC [**Figure 1A**]. Data were filtered to retain only variants private - i.e., found exclusively in just one population - Non-Finnish European (NFE), African American (AFR), Admixed American (AMR), East Asian (EAS) or South Asian (SAS). The intent was to enrich for variants more likely to be ancestry-specific and thus better reflect factors influencing variant annotation for each selected population. These five populations cover diverse ancestries represented by >22,000 individuals each in gnomAD v4.0 [**Table S1]**. We used population assignments made by the gnomAD consortium based on principal component analysis of ancestry-informative markers (Karczewski et al., 2020). The number of private, protein-coding variants per population ranged from 773 (AMR) to 8011 (NFE) [**Figure 1B**], with the high number for NFE reflecting how NFE individuals make up 77% of samples in gnomAD. All populations have a minimum of 25 protein-coding variants in the genes of interest [**Table S2A**]. *BRCA2* and *ATM* had the most private protein-coding variants, with 3497 and 3063, respectively, while *TP53, RAD51C* and *RAD51D* had the fewest (431, 462 and 492) [**Table S3**].

**Figure 1:**
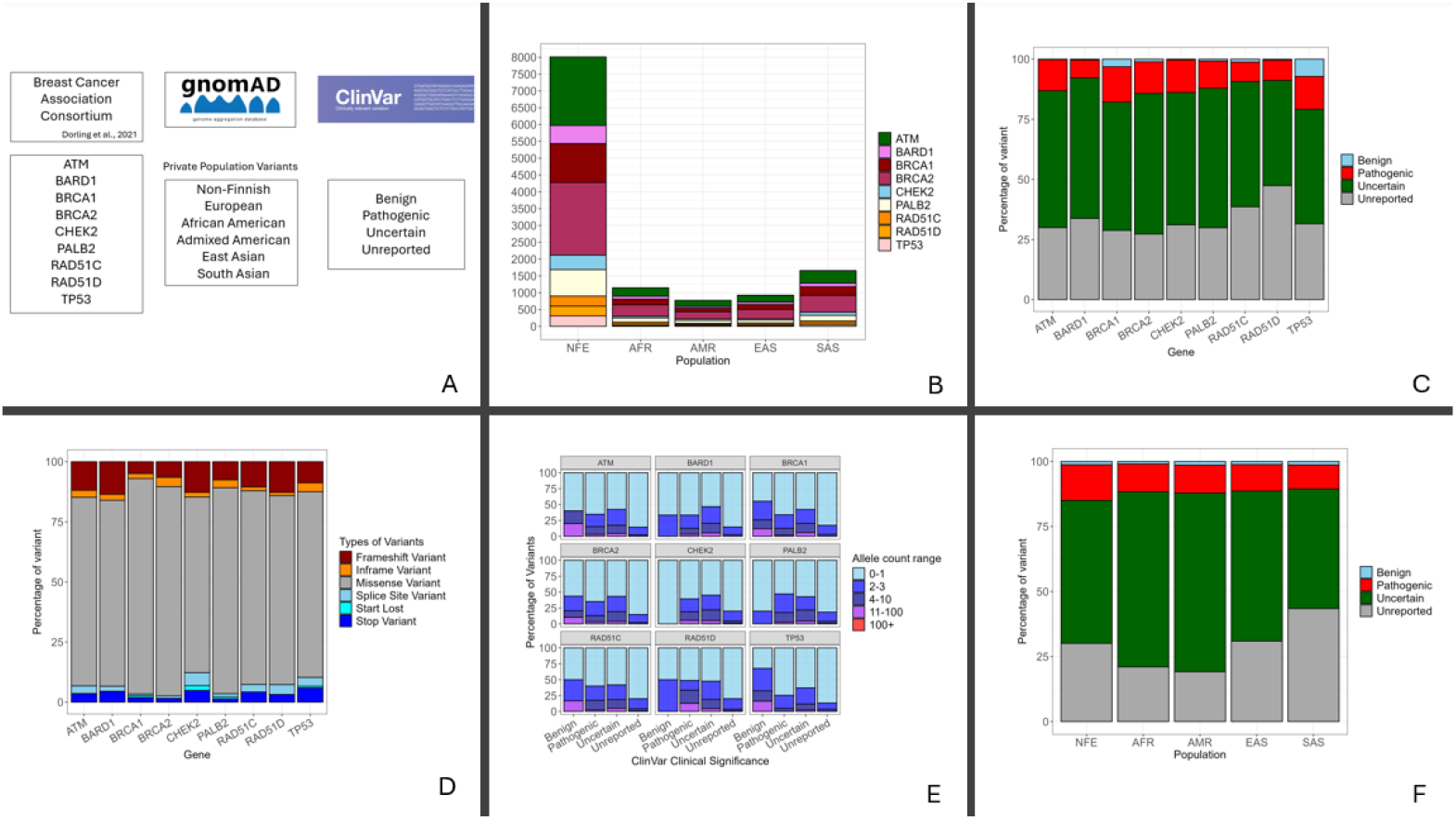
Features of protein-coding variants in breast cancer risk genes that are private to distinct populations in gnomAD. A – Study overview. Protein coding variants from nine genes identified by the Breast Cancer Association Consortium were selected from gnomAD for inclusion in the analysis if they were private to one of the populations shown and their ClinVar status as annotated in gnomAD were recorded. B – Total numbers of protein-coding variants from gnomADv4 private to each population, per gene. NFE - Non-Finnish European, AFR – African American, AMR - Admixed American, EAS - East Asian and SAS - South Asian. C – Percentages of population private variants within each ClinVar annotation category, per gene. The height of each segment indicating its percentage relative to the total number of variants for the corresponding gene. D – Percentages of population private variants with Unreported status by Ensemble VEP annotation across each gene. The height of each segment represents the percentage of the variant type within Unreported variants for the respective gene. E – Frequency ranges of population private protein-coding variants included in the analysis. Each bar displayes the percentage of variants with the indicated number of alternate alleles (0-1, 2-3, 4-10, 11-100, 100+) in gnomAD v4 in each gene for each ClinVar annotation category . F – Percentages of population private variants within each ClinVar annotation category, per populations. The height of each segment indicating its percentage relative to the total number of variants for the corresponding population.

We next considered the ClinVar annotation status as given in gnomAD for each variant in our filtered data set. Variants listed as ‘Unreported’ represent potential breast cancer risk SNVs and indels that did not have assessments of pathogenicity in ClinVar at the time data were gathered for this study (March 2024). Proportions of Unreported variants provide a comparative estimate of how rates of reporting of expert clinical curations vary across genes and populations.

Of the 12,520 variants included in our analysis, 3804 had ClinVar status of Unreported (30.4%), with *BRCA2* having the lowest rate of Unreported variants (27.3%) and *RAD51D* the highest rate (47.4%) [**Figure 1C**]. Overall, the majority of Unreported variants were annotated as missense variants (82.5%) ranging from 73.0% in CHEK2 to 87.0% in BRCA2. However, altogether 665 frameshift, splice site, start lost, stop gained and inframe deletion variants were Unreported across all genes, suggesting a substantial number of potentially pathogenic breast cancer risk variants are currently absent from ClinVar.

Unreported variants also tended to be rare, more often found in a single individual within a population than are variants reported in ClinVar. Across all genes, 80% or more of the Unreported variants were singletons, having an allele count of 1 in gnomAD v4 [**Fig 1E]**. In contrast, <65% of all Benign, Pathogenic or Uncertain variants reported in ClinVar had allele counts of 1 across our 9 genes of interest [**Table S4**]. Benign variants from *CHEK2* and *PALB2* were exceptions, but only 2 and 10 Benign variants were found in these genes, respectively. Singletons comprised 74.5% of the Pathogenic variants reported from TP53, a gene where germline coding variants are known to be under strong negative selection and usually rare (Kou et al., 2023).

Overall, substantial numbers of variants private to single populations and of undetermined clinical significance that are predicted to alter protein coding sequence of breast cancer risk genes could be found in gnomAD v4. Most were rare missense variants, although high-impact mutations such as frameshift, stop gained and splice site alterations accounted for 17.5% of Unreported variants. Notably >30% of private variants were not reported in ClinVar, representing potential missing information about the landscape of breast cancer risk across populations of diverse ancestry.

### Putative breast cancer risk variants from South Asian populations are less likely to be reported in ClinVar than variants from other populations

We assessed the impact of population ancestry on depth and availability of data for putative breast cancer risk variants in ClinVar by comparing the rates at which private, protein coding variants from gnomAD v4 across the five populations of interest were reported in ClinVar. The overall percentage of Unreported of all variants combined in our data set was 28.9%, but the rates at which population-specific variants were reported in ClinVar varied. AMR had the lowest rate of Unreported variants (19.1%), followed by AFR (21.0%), NFE (30.0%), and EAS (30.9%), while SAS had the highest rate (43.4%) [**Table S2B, Figure 1F**].

The SAS population had the lowest or among the lowest reporting rates for all 9 genes assessed [**Figure 2]**. The percentage of gnomAD variants in ClinVar was 5.3% to 19.0% lower for SAS than for NFE across all genes [**Table S2B]**. Accounting for total numbers of observed variants from gnomAD, the difference was nominally significant (unadjusted Fisher Exact p-value < 0.05) in 7 of the 9 genes, with *RAD51C* and *RAD51D* the exceptions, and exceeded the Bonferroni multiple hypothesis testing threshold (p < 0.0014) in 3 genes (PALB2, ATM, BRCA2). Further, results were highly significant (Fisher Exact p-value < 2.2e-16) when variants were combined across all genes. Variants from the gnomAD SAS population are thus consistently under-represented in ClinVar across a trend across established and putative breast cancer risk genes.

**Figure 2:**
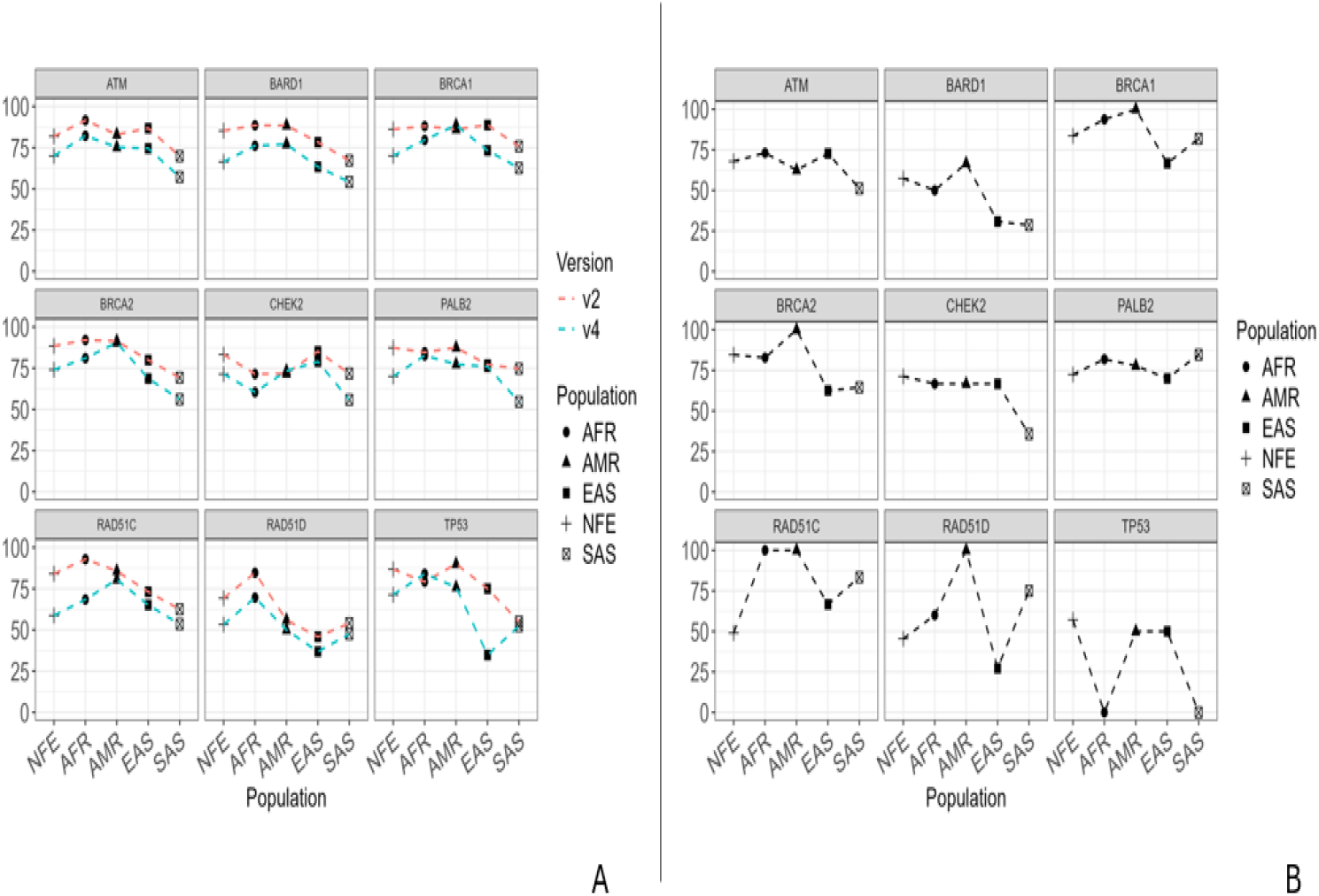
ClinVar reporting rates for protein-coding variants in breast cancer risk genes that are private to distinct populations in gnomAD. A – The percentage of variants private to each indicated gnomAD population that are reported in ClinVar, for each gene. Data from gnomAD v4 (Blue line) are overlaid onto data from gnomAD v2 (Red line). B – Percentage of private frameshift, stop gained and splice site variants from each gnomAD population that are reported in ClinVar, for each gene.

Notably, ClinVar reporting rates for private, protein coding variants from gnomAD were typically comparable between NFE, AFR, AMR and EAS **[Figure 2A]**. In fact, reporting rates were higher in AFR and AMR than in NFE for 8/9 genes, with unadjusted Fisher Exact p-value < 0.05 in 4 genes for AFR versus NFE, and in 3 genes for AMR versus NFE **[Table S5]**. Beyond the data from SAS, reporting rates substantially lower than those of NFE were only observed for EAS in RAD51D (36.8% vs 53.3%) and TP53 (34.8% vs 71.3%). Thus, in our analysis under-reporting of private, protein coding variants in breast cancer risk genes in ClinVar was much more prevalent in the population of South Asian ancestry.

As reported, the gnomAD v4 release had a 8-fold increase in the NFE population from gnomAD v2, from 77,165 in to 622,057, and a 3-fold increase in the SAS population, from 15,308 to 45,546. Adding more individuals would be expected to increase the numbers of population-private variants in gnomAD v4, perhaps without sufficient time to have elapsed for them to be expertly curated and deposited into ClinVar. To assess the extent to which the expansion of gnomAD may contribute to differences in ClinVar reporting rates between NFE, SAS and the other populations studied here, we repeated our analyses using data from gnomAD v2 [**Methods**].

While ClinVar reporting rates are lower for gnomAD v4 than gnomAD v2 in nearly every population and genes assessed, the relative differences between populations remained largely concordant, as indicated by the similar shapes to the curves in **Figure 2A**. The proportion of private, protein-coding variants reported in ClinVar was again consistently significantly lower across genes in SAS relative to NFE (unadjusted Fisher Exact p-value < 0.05 for 7/9 genes) **[Table S6]**. Reduced ClinVar reporting rates for putative breast cancer risk variants in the SAS population was therefore a trend also present in gnomAD v2.

The majority of Unreported variants across all genes and populations (82.8%) are annotated as causing non-synonymous (missense) changes [**Table S7**]. However, variants that truncate proteins of alter exon structure are likely to be more amenable for inclusion in ClinVar, given their more profound and readily interpretable effects on protein function. To assess how possible biases toward reporting more impactful variants in ClinVar may influence our findings, we compared combined reporting rates for frameshift, stop gained, splice site and inframe deletion variants from gnomAD v4 between populations. ClinVar reporting rates were lower in SAS than NFE and other populations for 6 of the 9 genes investigated **[Figure 2B]**. The comparison between SAS and NFE attaining a nominal Fisher Exact p-value < 0.05 for *ATM, BRCA2, CHEK2* and *TP53* [**Table S8**]. As expected, ClinVar reporting rates for missense variants was significantly lower for SAS across all genes [**Figure S1**]. These data show under-reporting of putative risk variants from the SAS population occurs for multiple types of protein-altering variants.

Further, we see little difference in the proportion of variants in ClinVar that are labelled Pathogenic for each population [**Table S8**]. If anything, SAS variants are slightly more likely to be labelled Pathogenic in ClinVar than for the other populations (unadjusted Fisher Exact p-value = 0.04; Odds Ratio = 1.2). Thus, perceptions that variants from SAS populations are less likely to be pathogenic and therefore less worthy of submission to ClinVar are unlikely to explain the under-reporting of putative breast cancer risk variants from SAS individuals.

ClinVar is considered diverse and comprehensive, but may still lack variants found in other databases. However, only 12 of the Unreported population private gnomAD in our study could be found in an independent database of annotated variants from cancer risk genes, FLOSSIES (Walsh et al., 2010) [**Table S9**]. The majority were from the NFE and AFR populations, with just 2 from SAS. Overall reporting rates of gnomAD variants in for FLOSSIES were quite low, in keeping with its smaller size. However, reporting rates were still higher for NFE and AFR than for SAS, EAS and AMR across all genes [**Figure S2**]. This does reflect the composition of FLOSSIES but also demonstrates how variants from SAS and other non-European populations may be under-represented in variant annotation databases generally.

To investigate whether Unreported germline variants may have been identified and characterized in tumors, we searched for them in OncoKB, which curates functional evidence for pathogenicity of recorded somatic mutations (Chakravarty et al., 2017). Generally, <20% of variants that were Unreported in ClinVar had pathogenicity assessments in OncoKB, across each gene and each population [**Figure S3A**]. This was especially pronounced for missense variants, where <10% were assessed by OncoKB [**Figure S3B**]. The exception was TP53 where every population except AMR had reporting rates of above 25% for all populations, even for missense variants. However, functional information is still unavailable for the majority of population-private Unreported variants identified in our study.

### Computational analysis of pathogenicity assessment reveals Unreported missense variants with potential impacts on breast cancer risk

While protein-truncating, frameshift and canonical splice site variants have clear links to pathogenicity, the majority of Unreported variants are missense, which vary in their functional effects. To evaluate the extent to which Unreported missense variants may contribute to breast cancer risk, we extracted pathogenicity predictions from dbNSFP (Liu et al., 2020) for all population-private gnomAD missense variants in our 9 genes of interest [Methods]. Rank scores scaling pathogenicity predictions from 0 to 1 were utilized for each algorithm, which allows direct and meaningful comparison across algorithms and ClinVar annotation categories.

As illustrated by rank scores based on the Combined Annotation Dependent Depletion (CADD) algorithm (Rentzsch et al., 2019), variants annotated as Pathogenic in ClinVar tend to have rank scores at or close to 1, while variants annotated as Benign have much lower rank scores, close to 0 [**Figure 3A**]. Rank scores for variants categorized as having Uncertain pathogenicity in ClinVar were distributed broadly from 0 to 1 consistent with a range of possible effects for variants with this designation. Variants in the Unreported category showed a similar range of CADD rank scores as Unreported variants, indicating they may have variable effects on protein function too. Similar patterns were observed for rank scores derived from AlphaFold predictions (Jumper et al., 2021) from missense variants [**Figure 3B**], as well as other algorithms included in dbNSFP [**Figure S4**].

**Figure 3:**
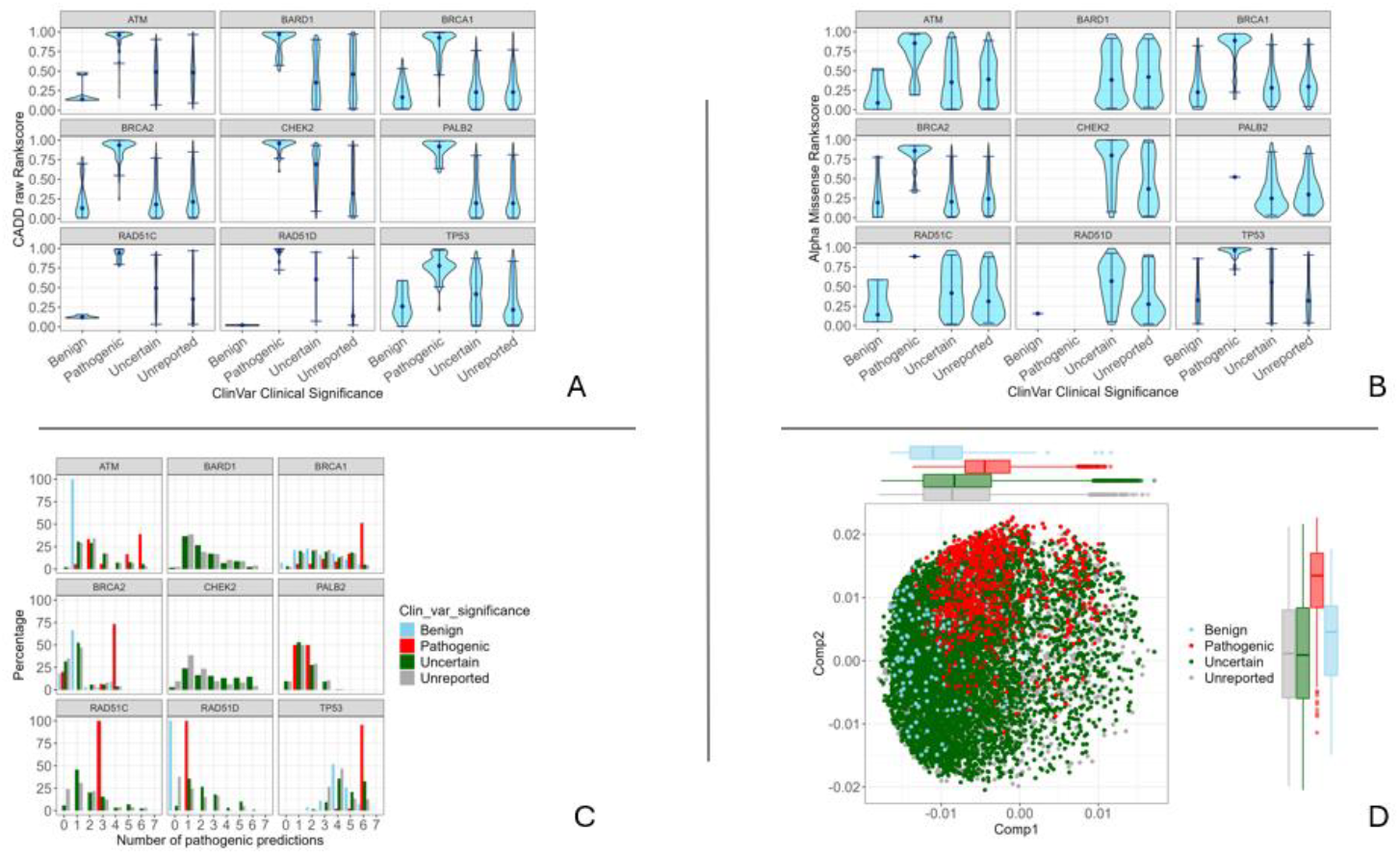
*In silico* prediction of pathogenicity for non-synonymous variants in breast cancer risk genes that are private to distinct populations in gnomAD. A – Violin plots of raw CADD rankscores from dbNSFP of gnomAD variants private to each indicated population, divided by gene and ClinVar annotation category. The widths of the violin represent the density of variants, with the individual points marking their medians. B – Violin plots of AlphaMissense rankscores from dbNSFP of gnomAD variants private to each indicated population, divided by gene and ClinVar annotation category. C –The number of pathogenicity prediction algorithms returning indicating likely pathogenicity for gnomAD variants private to NFE, AFR, AMR, EAS or SAS that are listed as Pathogenic (red), Benign (blue), Uncertain significance (green) or Unreported (grey) in ClinVar, in each gene. Results shown are for variants from all populations combined. D – Clustering by principal component analysis (PCA) of gnomAD variants private to NFE, AFR, AMR, EAS or SAS across genes, color-coded by ClinVar annotations based on 24 rankscores of protein pathogenicity prediction algorithms from dbNSFP. Points represent individual variants, coloured by ClinVar annotation category. The first two principal components for each annotation category are displayed in the plot margins. Results shown are for variants from all populations and all genes combined.

We assessed consistency in predictions of variant impacts by aggregating data from six algorithms in dbNSFP plus AlphaMissense (Cheng et al., 2023) that provide an assessment of whether each population private missense variant was either damaging/deleterious or tolerated/benign (PolyPhen, SIFT, Mutation Assessor, FATHMM, MetaSVM, MetaLR and AlphaMissense), from variants for which predictions from at least 4 of these algorithms were available [Methods]. Generally were predicted to be damaging by 0 or 1 of the algorithms Overall, 40.17% of variants classified as Benign in Clinvar included in our study were predicted damaging by 1 or fewer algorithms, with multiple exceptions seen in BRCA1 and TP53 [**Figure 3C**]. Conversely, a high percentage (78.95%) of variants labelled Pathogenic in ClinVar were in our data set were predicted to be damaging by a majority of (4/7) algorithms, in line with expectations. Once again Uncertain and Unreported variants showed a range of results in the number of algorithms that predicted damaging effects [**Figure 3C**].

To comprehensively compare the potential functional impacts of Unreported missense variants across genes and populations, Principal Component Analysis (PCA) was used to cluster and visualise differences between 9,827 variants across all genes [**Table S10**] based on their rankscores from 24 predictive algorithms [**Table S11**; Methods]. Separation of Benign and Pathogenic variants was observed while Uncertain and Unreported variants again showed a wider range of values across the first two principal components [**Figure 3D**]. Collectively, predictions of variant pathogenicity indicate a subset of population private missense variants not yet reported in ClinVar may be capable of causing deleterious changes to protein function.

## Discussion

The depth of knowledge of cancer genomics varies between populations according to genetic ancestry. This study investigated how this affects current data on genetic risk factors for breast cancer in people of European, African, American, East Asian and South Asian ancestry. Proportions of private variants reported in ClinVar in 9 recognized breast cancer risk genes were similar between NFE, AMR, AFR and EAS populations in gnomAD. Variants in breast cancer genes from these populations may be reported to ClinVar at comparable rates, acknowledging the over-representation of NFE in genomic data. Where differences were found, AMR, AFR and/or EAS often had higher proportions of private variants in ClinVar, likely related to the broader sampling of NFE resulting in larger numbers of private variants.

In contrast, lower percentages of protein-altering variants private to the SAS population were reported in ClinVar than NFE and other populations for all genes examined here. This represents a deficit in available knowledge of genetic risk factors for breast cancer in South Asians. The differences were statistically significant across a majority of genes and observed across variant types and in the, FLOSSIES database too.

Practical and cultural barriers to genetic screening and cancer treatment in South Asian communities may account for lower reporting rates for SAS variants in ClinVar (Allford et al., 2014; Hann et al., 2017) in addition to poor information exchange between data repositories from different parts of the world (Rehm et al., 2021; Wright et al., 2019). Although lifetime risk of cancer is thought to be lower in South Asians (Gathani et al., 2014; Tran et al., 2018), as one of the world’s largest ancestry groups, their overall burden of cancer is substantial (Jain et al., 2024; Pramesh et al., 2022) and increasing due to lifestyle changes (Mubarik et al., 2022).

Many Unreported variants could be labelled variants of uncertain significance (VUS) by current American College of Medical Genetics criteria (Edsjö et al., 2023; Yadav & Couch, 2019), perhaps reducing likelihood of reporting. However, our analyses reveal 9% of Unreported variants cause frameshift mutations (Table S7), while numerous others show evidence of potential functional effects (Figures 1 & 3). Such data warrant efforts to facilitate expert evaluation and reporting of putative pathogenic variants from South Asians and others with ancestry outside Europe.

It is widely known historical biases have left substantial gaps in our knowledge of cancer in populations with non-European ancestry (AACR CANCER DISPARITIES PROGRESS REPORT 2024, 2024). However, ways to identify and characterise such gaps are lacking. Our framework therefore creates an important methodological advance that brings to light shortfalls in publicly accessible data on breast cancer risk variants from South Asian populations. Overall, South Asians remain poorly represented in cancer genomics research (Pramesh et al., 2022; Thorn et al., 2024) and our results indicate one of the potential consequences of this shortfall. Our work motivates deeper study of the genetics of cancer in this ancestry group and efforts to identify knowledge gaps between ancestries in other areas of cancer genomics. Whether similar deficits in knowledge of risk variants for other human diseases exist for South Asian populations warrants further consideration as well.

## Data Availability

Code and data files used in these analyses are available as Git repo at https://github.com/RaveenRony/ClinVar-annotation-rates

## Acknowledgments

We wish to acknowledge the people of the Kulin Nations, on whose land these studies were done. We pay our respects to their Elders, past, and present. Dr. Goode received support from the Victorian Cancer Agency (MCRF17005) and the Peter MacCallum Cancer Foundation. Dr Doig received support from the Laby Foundation. We thank the Peter MacCallum Centre Research Computing Facility for technical support, as well as Dr Justine Marum, Dr Sally Hunter and A/Prof Mahesh Iddawela for helpful feedback and suggestions.

## Author contributions

DLG conceived and directed the study. RR, SD and SY sourced data, wrote code and performed analyses. KD provided variant annotations and pathogenicity predictions and advised on analyses. RR and DLG wrote the manuscript. All authors reviewed and revised the manuscript and approved the final version.

