## Supplementary Figures for "Putative breast cancer risk variants from populations of South Asian ancestry are under-represented in public variant classification databases"

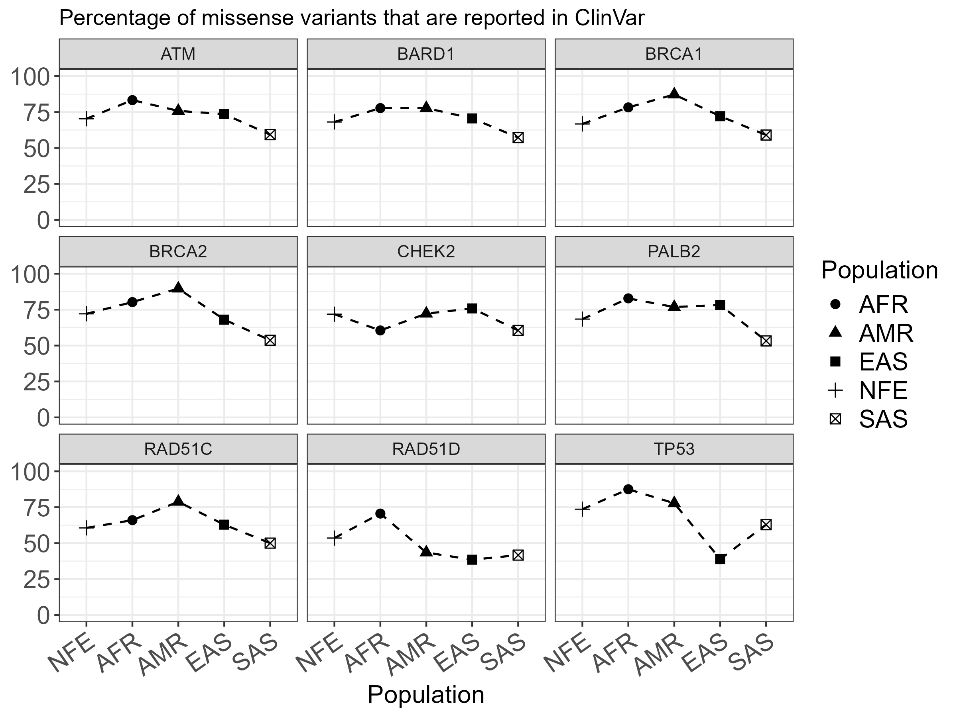
Fig S1 – Reporting rates of missense variants from GnomAD

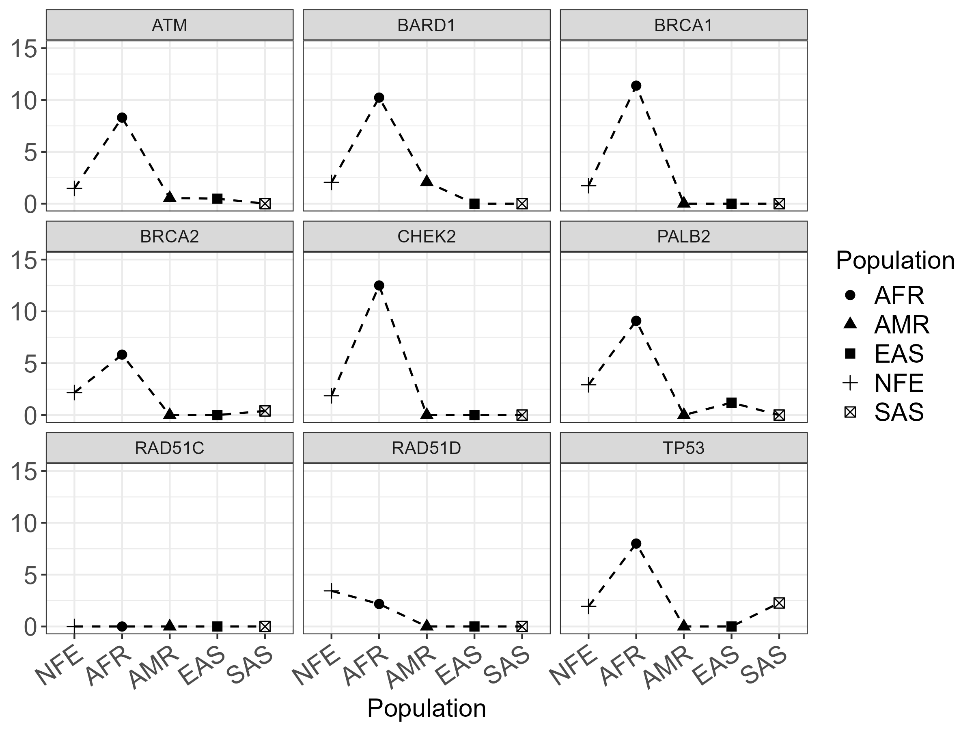
Fig S2 – Reporting rates of variants from GnomAD in the FLOSSIES database.

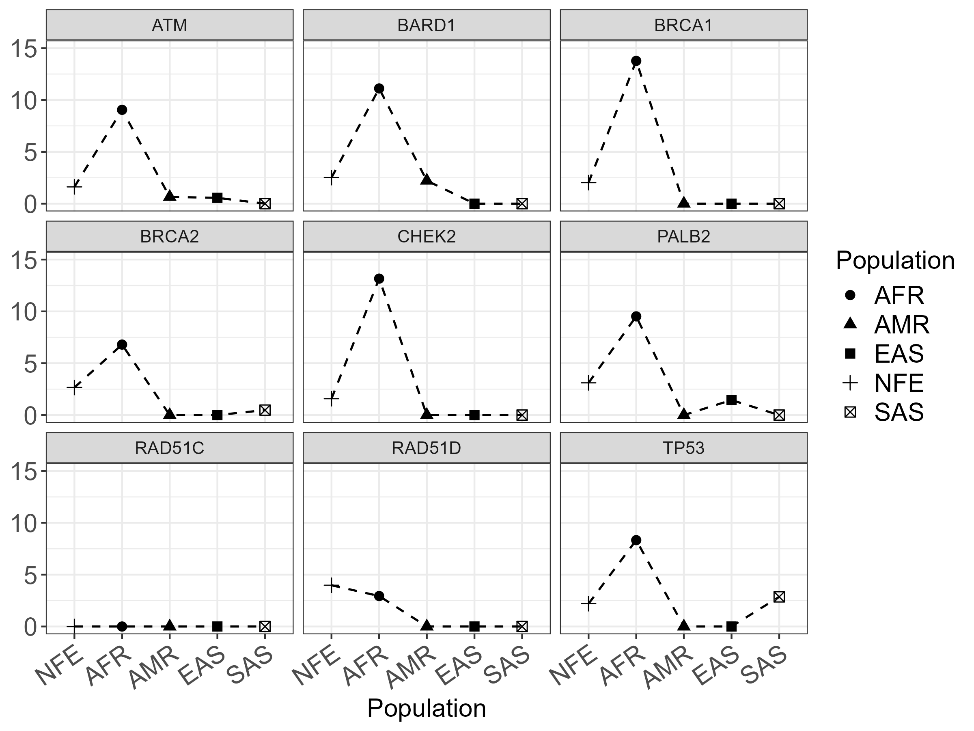

Fig S2A - Reporting rates of frameshift, inframe, splice site and stop gained variants from GnomAD in the FLOSSIES database.

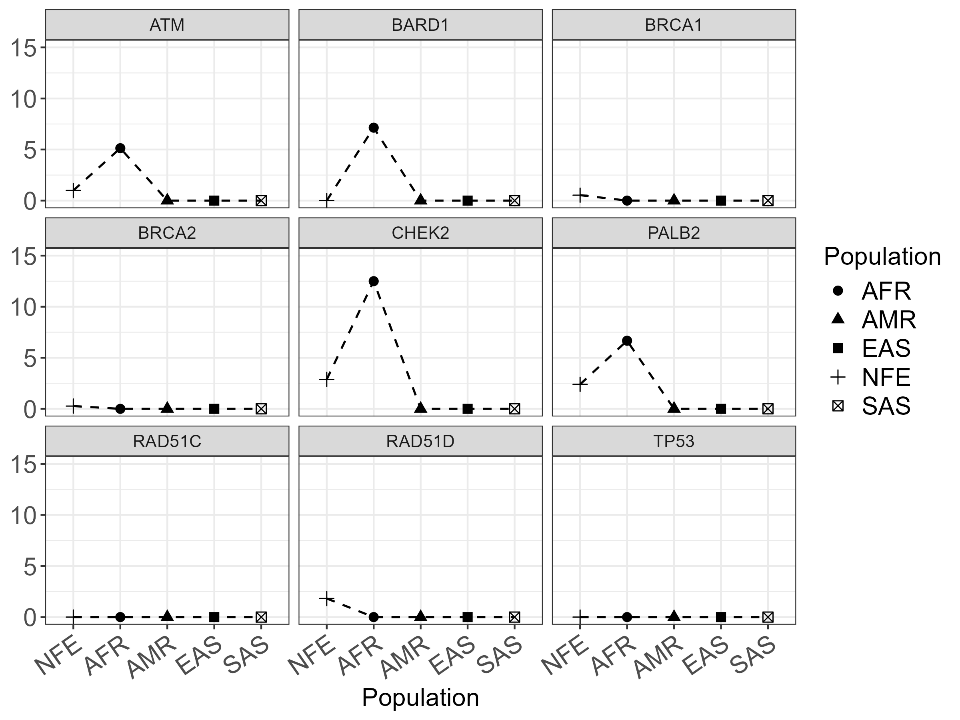
Fig S2B – Reporting rates of missense variants from GnomAD in the FLOSSIES database.

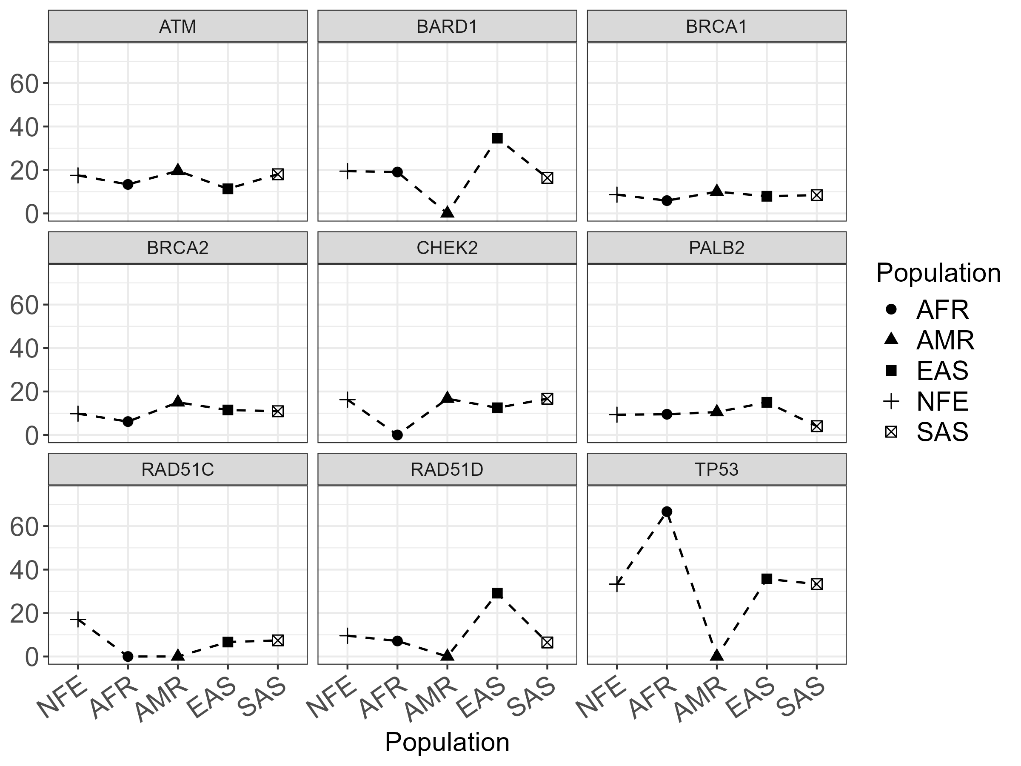
Fig S3 – Reporting rates of variants from GnomAD in the OncoKB database.

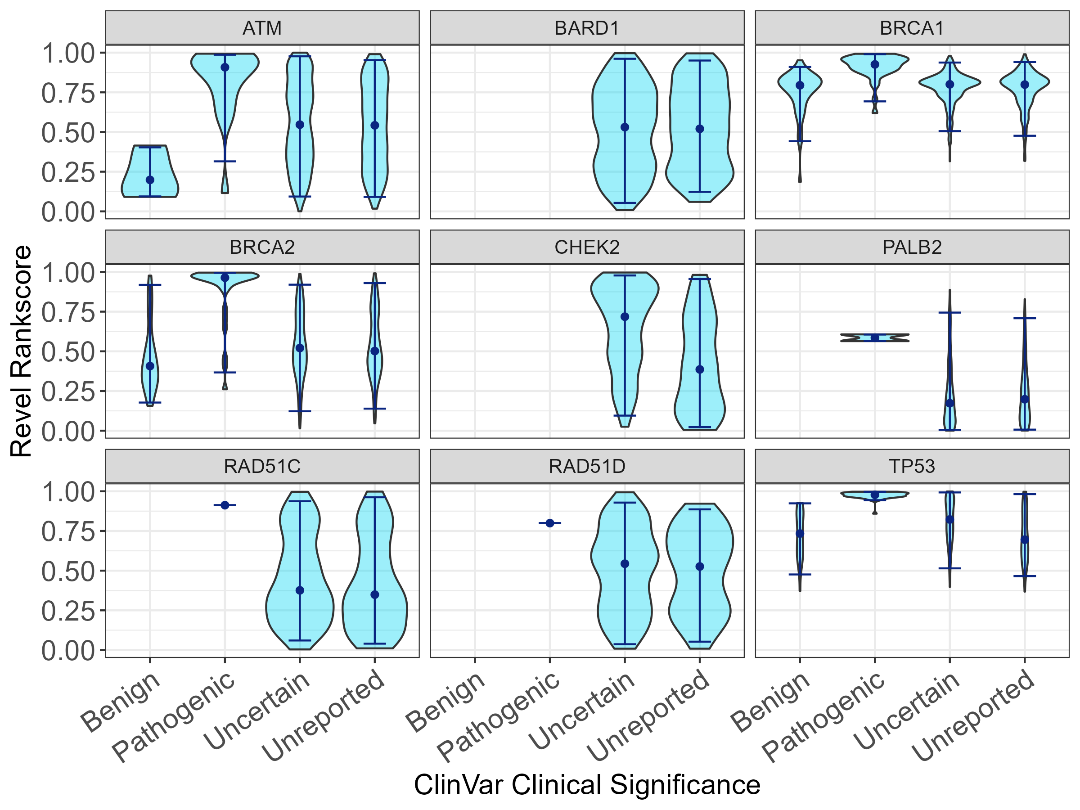
Fig S4 A – Violin plots of Revel rankscores.

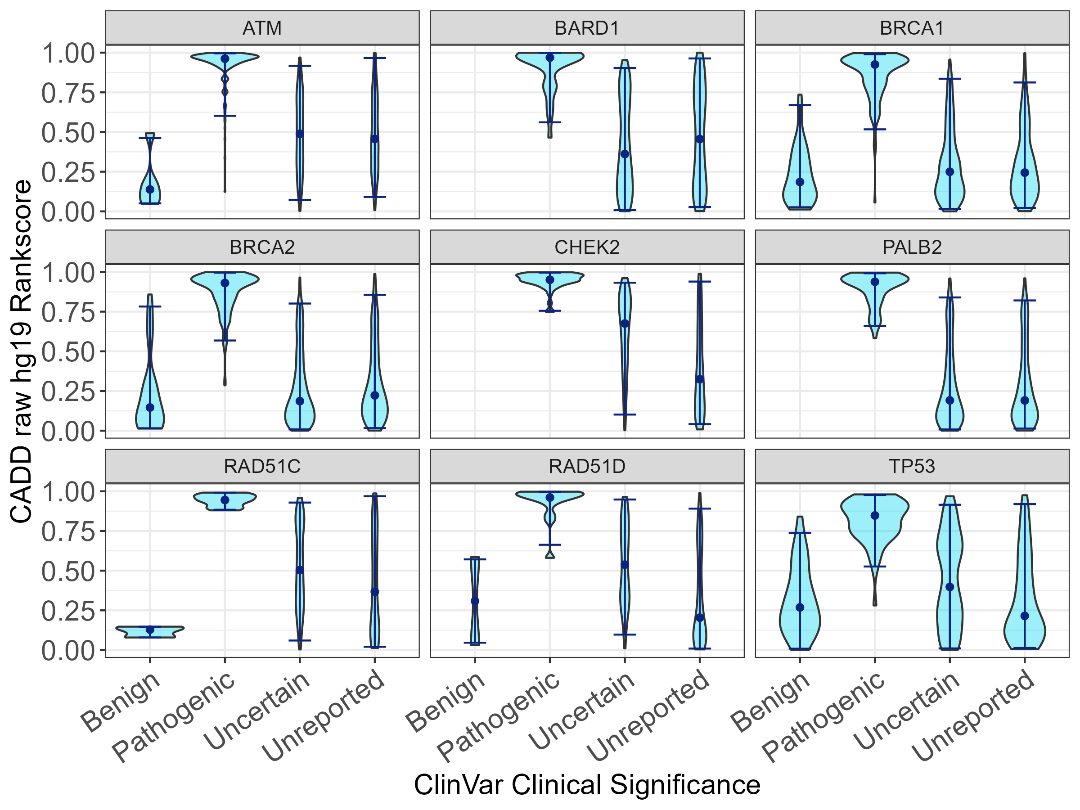
Fig S2B – Violin plots of CADD raw hg19 rankscores.

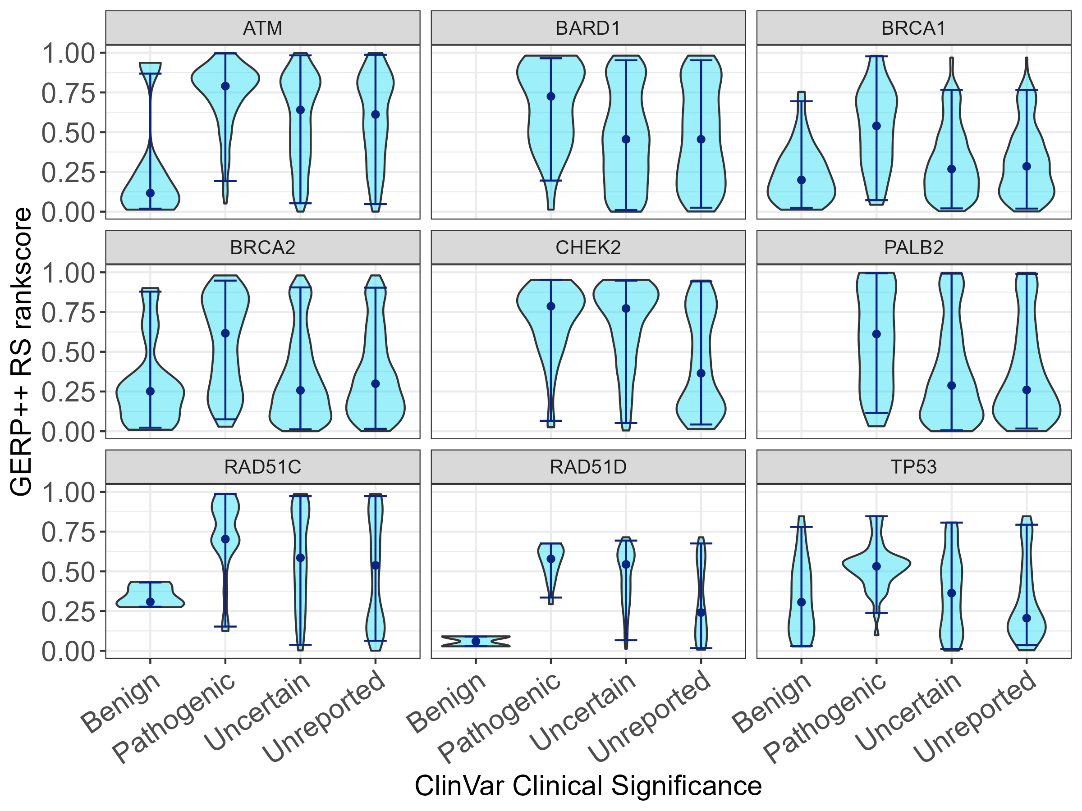
Fig S4C – Violin plots of GERP++ RS rankscores.

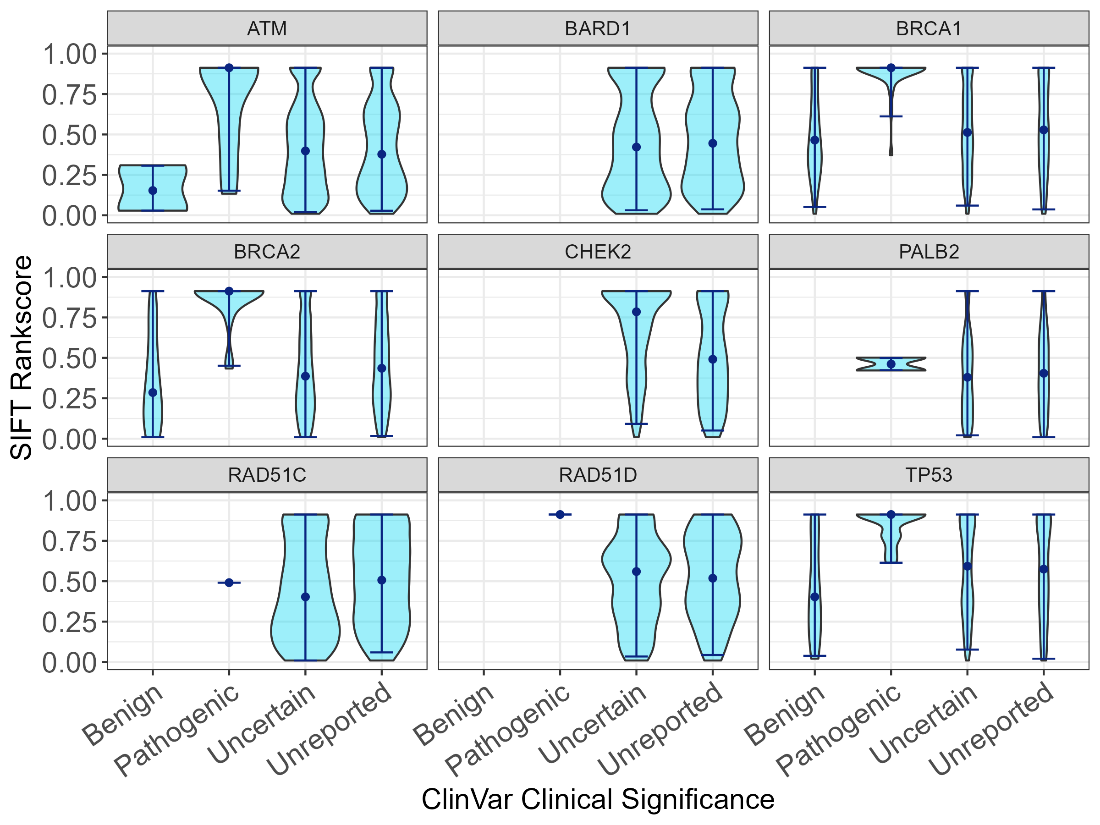
Fig S4D – Violin plots of SIFT rankscores.

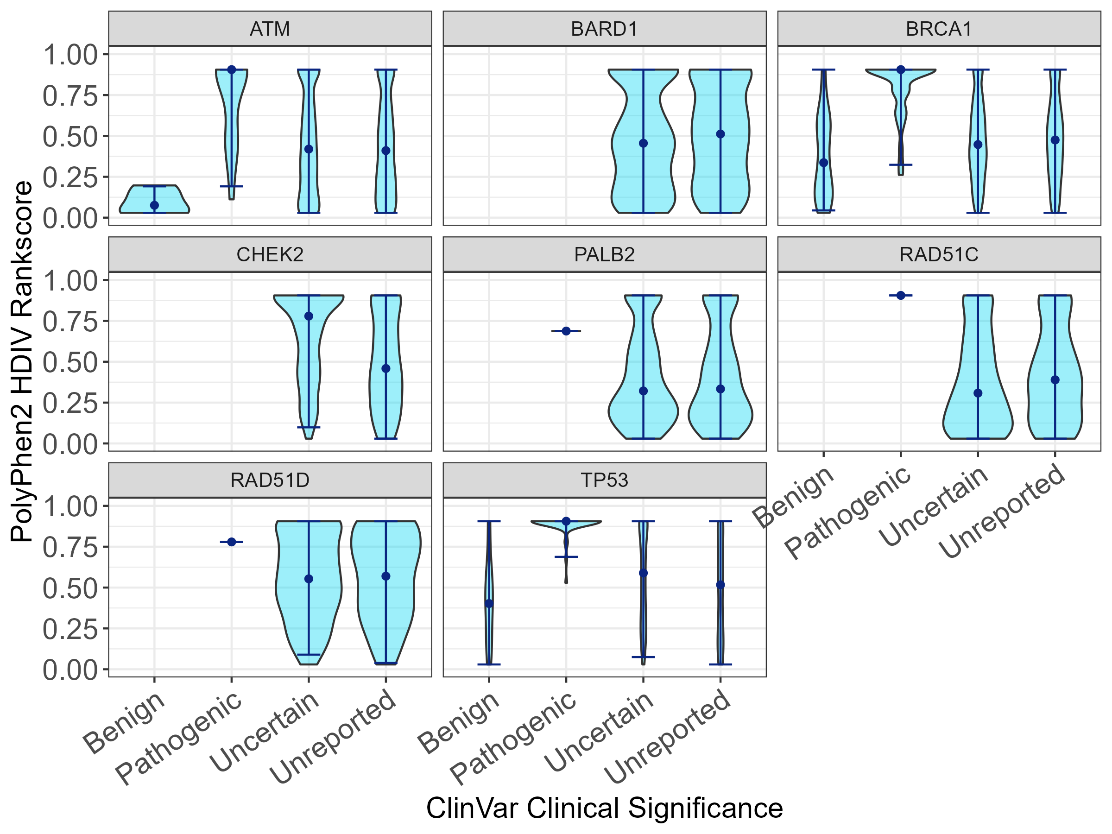
Fig S4E – Violin plots of PolyPhen2 HDIV rankscores.

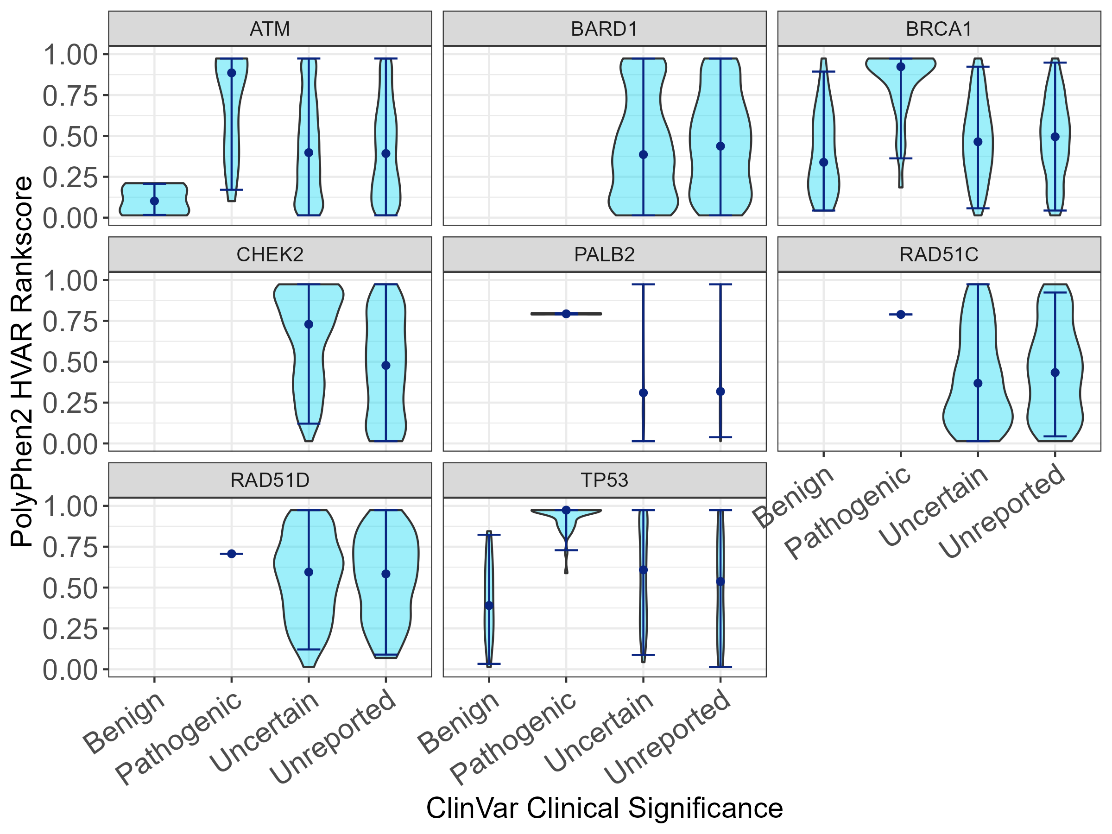
Fig S4F – Violin plots of PolyPhen2 HVAR rankscores.

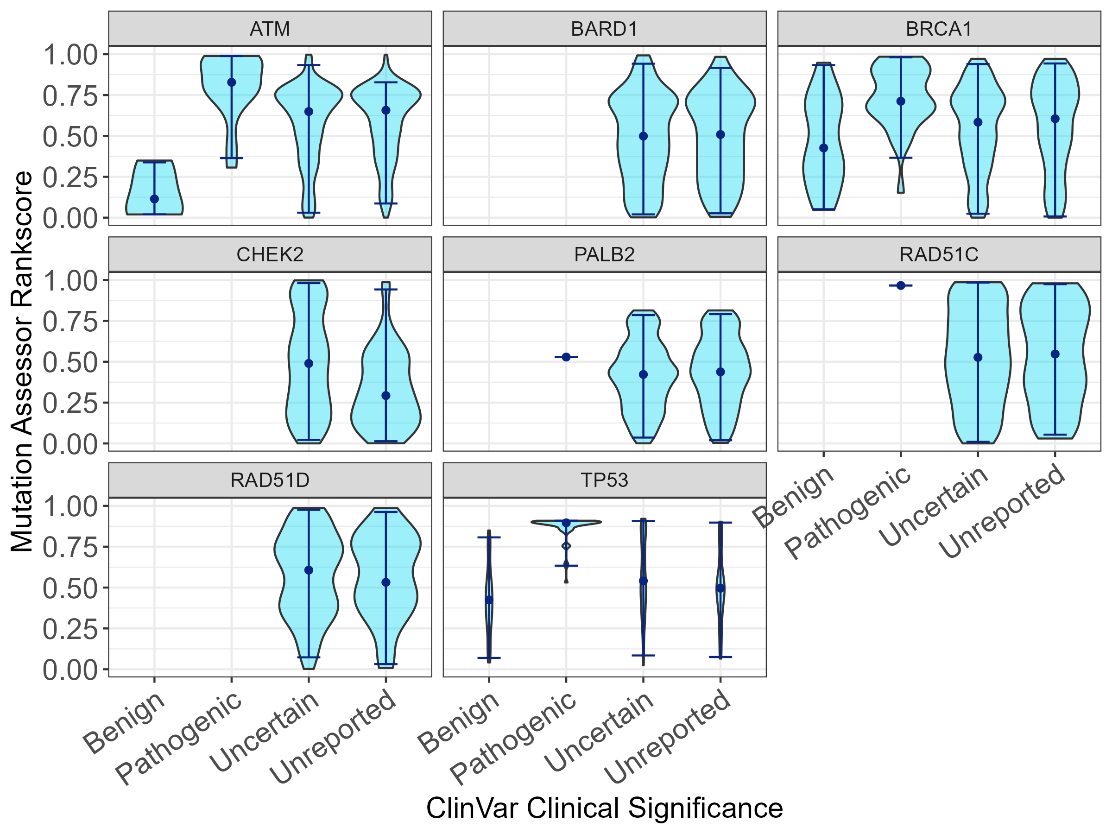
Fig S4G – Violin plots of Mutation Assessor rankscores.

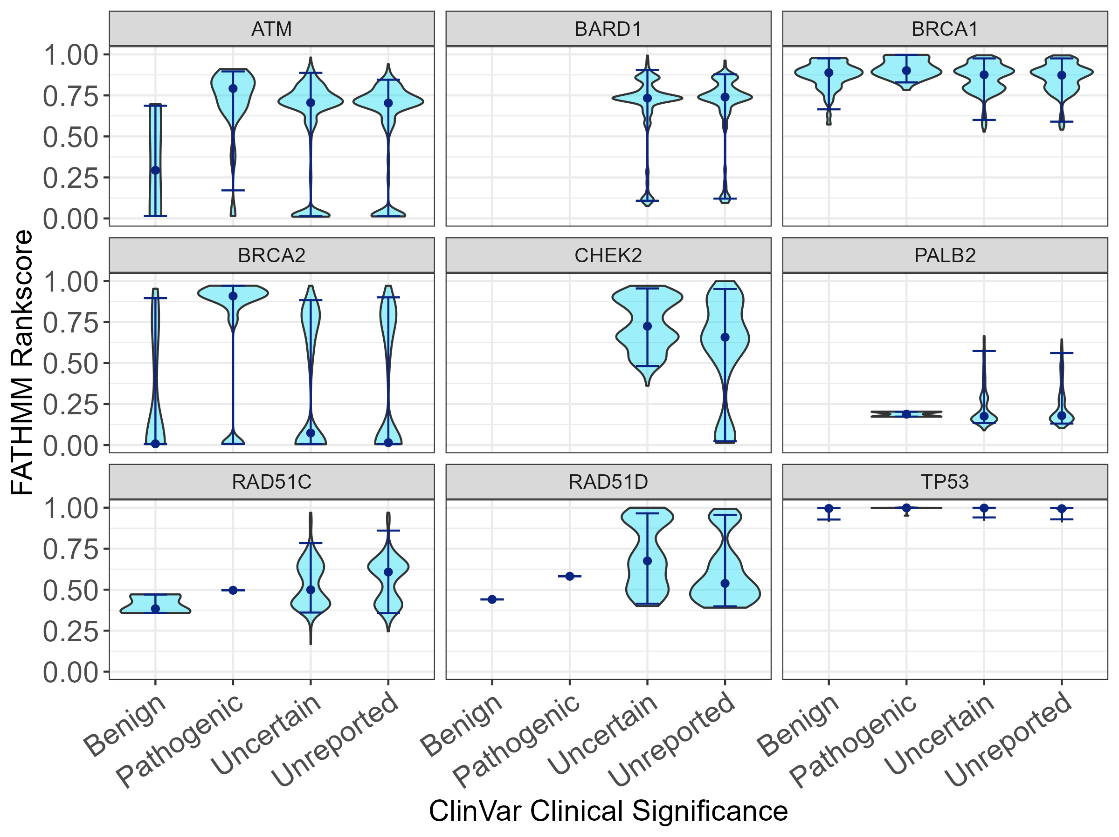
Fig S4H – Violin plots of FATHMM rankscores.

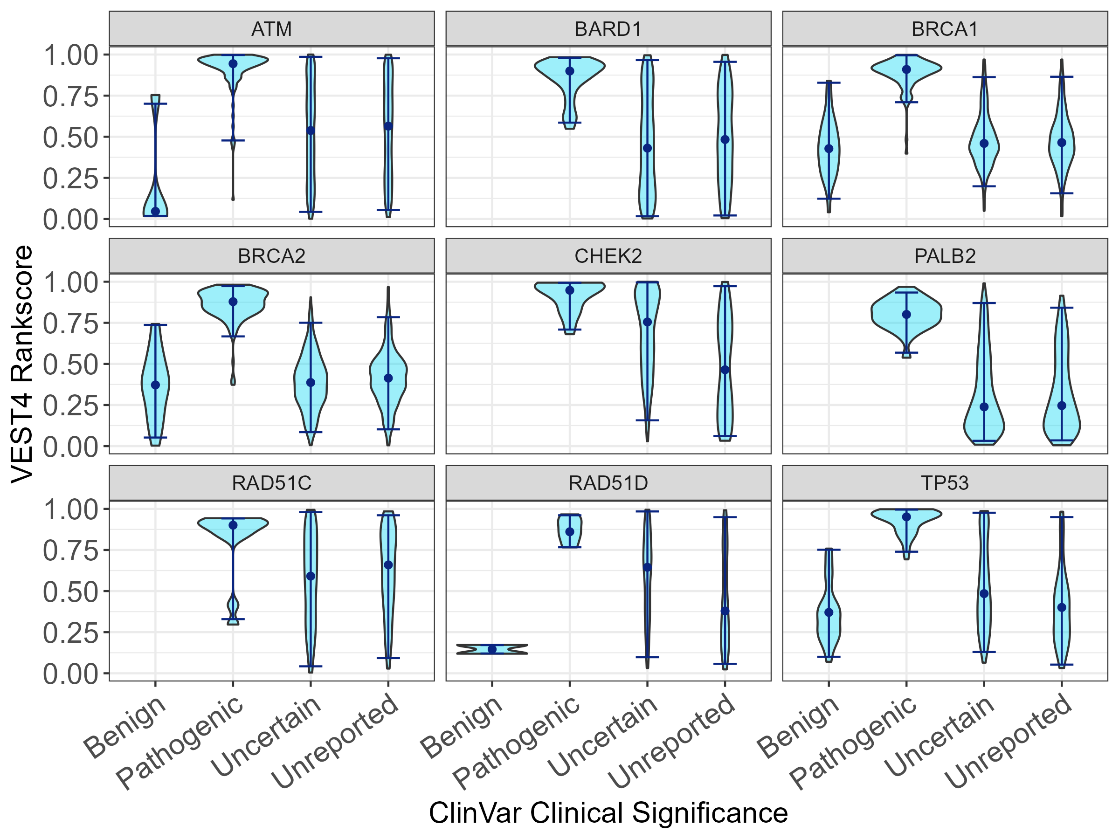
Fig S4I – Violin plots of VEST4 rankscores.

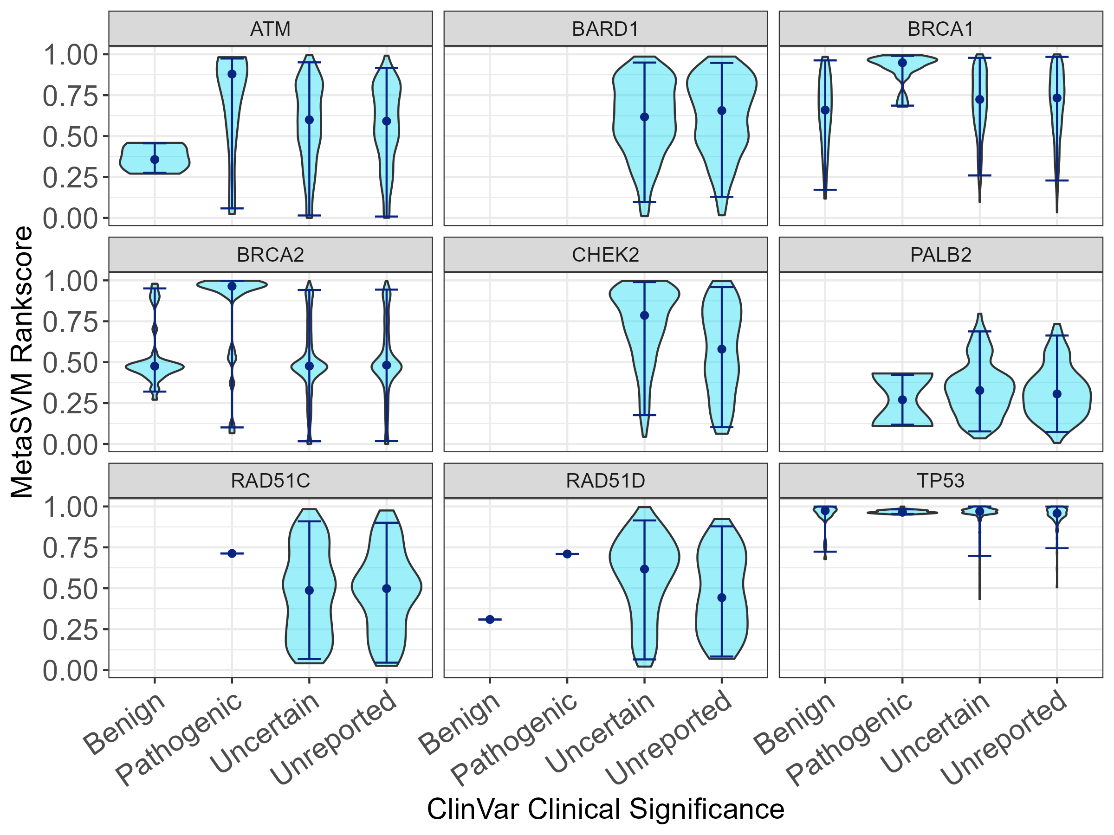
Fig S4J – Violin plots of MetaSVM rankscores.

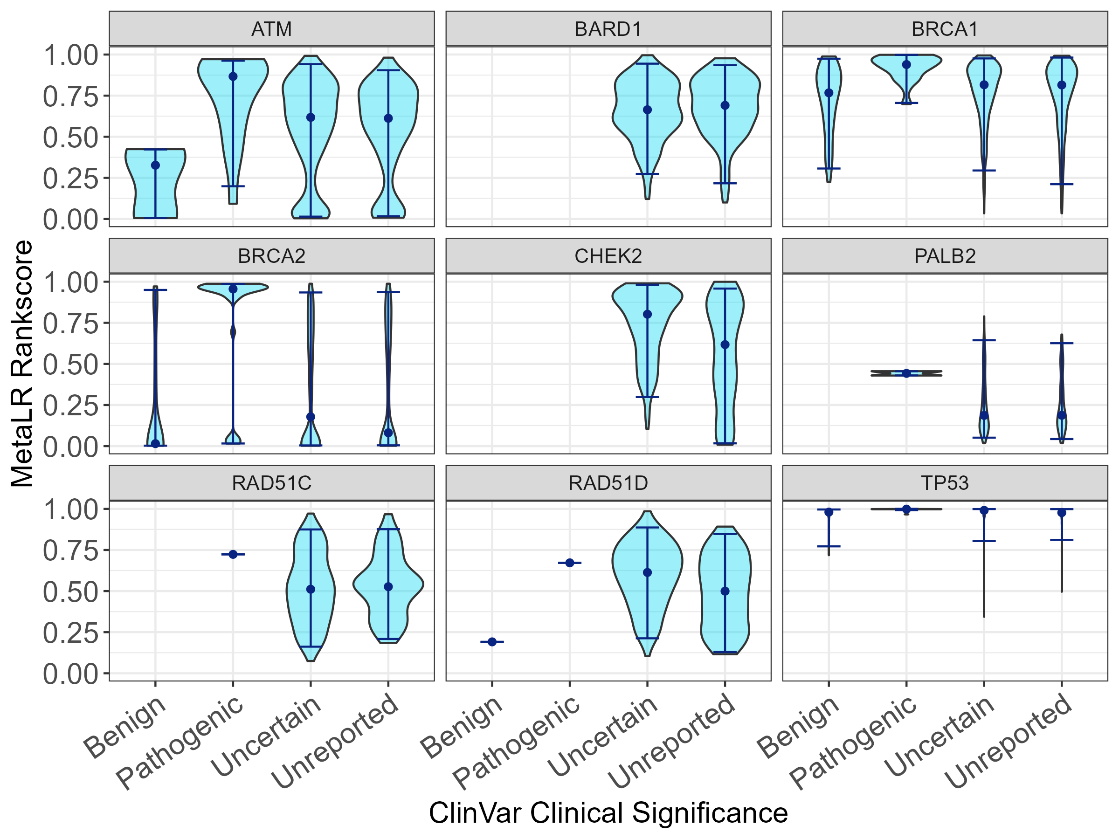
Fig S4K – Violin plots of MetaLR rankscores.
